# Pruning Digital Contact Networks for Meso-scale Epidemic Surveillance Using Foursquare Data

**DOI:** 10.1101/2021.09.29.21264175

**Authors:** Sofia Hurtado, Radu Marculescu, Justin A. Drake, Ravi Srinivasan

## Abstract

With the recent boom in human sensing, the push to incorporate human mobility tracking with epidemic modeling highlights the lack of groundwork at the meso-scale (e.g., city-level) for both contact tracing and transmission dynamics. Although GPS data has been used to study city-level outbreaks, current approaches fail to capture the path of infection at the individual level. Consequently, in this paper, we extend the usefulness of epidemics prediction from estimating the size of an outbreak at the population level to estimating the individuals who may likely get infected within a finite period of time. To this end, we propose a network-based method to first build and then prune the dynamic contact networks for recurring interactions; these networks can serve as the backbone topology for mechanistic epidemics modeling. We test our method using Foursquare’s Points of Interest (POI) smart-phone geolocation data from over 1.3 million devices and show that we can recreate the COVID-19 infection curves for two major (yet very different) US cities (i.e., Austin and New York City) while maintaining the granularity of individual transmissions and reducing model uncertainty. Our method provides a foundation for building a disease prediction framework at the meso-scale that can help both policy makers and individuals of their estimated state of health and help with pandemic planning.

## I. Introduction

Epidemiological contact tracing is a form of non-pharmaceutical intervention that can help mitigate the spread of infectious diseases such as HIV, Tuberculosis, Ebola, and SARS [1]. During an outbreak, each new infection is meticulously investigated by a team of trained health officials to swiftly quarantine contacts made during a patient’s contagious period. This method of outbreak mitigation is typically effective in diseases that have visible and fast-moving symptoms. However, such approaches can be easily outrun by a virus that has high asymptomatic rates, ambiguous symptoms, and long contagious periods as seen with the SARS-CoV-2 virus [2].

The catastrophic speed at which COVID-19 propagated into a pandemic reveals the need to automate contact tracing on a large scale, given the interconnected world we live in nowadays. Stimulated by intercontinental travel and growth of densely populated cities, an outbreak like COVID-19 can propagate along the complex networks of people much faster today than, for example, in the 1400s where the highly infectious Black Plague traveled on average 1.5 km per day [3]. Building off this need to meet the challenges posed by the increasingly interconnected world, we can combine digital contact tracing with epidemic modeling to accomplish both charting the path of infection, as well as identifying those in need of quarantining.

In the aftermath of the COVID-19 pandemic, both research and industrial communities face the challenge to build an epidemiological surveillance infrastructure to inform policy makers of impending outbreaks. Though a wide range of wearable devices currently allow for individual mobility tracing [4-6], it remains unclear whether agent-based epidemic models can be directly applied to largescale mobility datasets without introducing significant uncertainty in predicting the number of infections [7]. In addition, the longevity of the current pandemic presents a challenge to policy makers in being able to accurately monitor both early and ongoing outbreaks at the meso-scale (e.g., city level).

Although contact tracing was meant to stop viral transmission at the micro-level (from one individual to another), and mathematical modeling was developed to understand disease dynamics at the macro-level, we aim to bridge the gap by simulating an outbreak at the city-level using real interactions from large populations. Our method narrows the uncertainty in epidemic modeling while maintaining the granularity of interactions between individuals.

In this paper we present three novel contributions:

1. We propose a new methodology for building co-location networks as a proxy for city-level digital contact tracing and illustrate its significance using Foursquare geo-mobility data (FS).
2. We identify consistent (i.e., meaningful, or non-volatile) interactions among people that can serve as the potential paths of infection across cities like Austin and New York City (NYC).
3. We show that applying a mechanistic epidemic model on stably recurring interaction not only leads to more accurate predictions than using the raw contact networks, but also decreases the model uncertainty significantly (as much as 95% based on FS data).

Taken together, our contributions provide a foundation for utilizing highly granular digital contact tracing to model epidemics at the meso-scale. The remainder of this paper is organized as follows: Section II presents some relevant prior work; Section III describes our approach, while Section IV presents our preliminary results. Finally, Section V concludes the paper and provides a discussion of future work.

## II. Previous Work

The ongoing pandemic catalyzed the use of human mobility data to predict the spatiotemporal spread of COVID-19 at the individual level. However, until recently, researchers focused largely on the macro- and micro-dynamics in both human mobility and epidemics. Next, we provide a summary of relevant prior work in these both directions.

### A. Mobility Modeling

Human mobility research has focused thus far on characterizing trajectories at the population (e.g., region, country) and individual level— however, less is known about collective behavior at the meso-scale (e.g., city, neighborhood). While observed in bank note trajectories, bills are found to traverse short distances more often than large distances [8]. To test that finding at the city level using GPS data, researchers discover that trip distances exhibit a power law distribution across many cities [9]. Furthermore, it has been found that trajectories maintain a routine radius of gyration between stops, thus indicating that people tend to frequent certain locations with regularity [10]. Of note, the Universal Visitation Law of Human Mobility formally identifies that the regularity of which people visit POIs also follows a power law distribution suggesting people favor some places heavily over others [11].

Research at the microlevel of human mobility largely covers next location prediction, as well as pedestrian dynamics. In fact, we not only understand how pedestrians move in crowds and navigate indoor locations for short periods of time [12,13], but also how pedestrians respond to the COVID-19 social distancing mandates [14].

These prior works are important in characterizing the predictability of an individual’s trajectory; however, we are now prompted by the COVID-19 pandemic to analyze mobility behavior of pairs (or small communities) of people, in spaces covering neighborhoods or cities. Because the call for social distancing prompted the formation of quarantine bubbles, our work aims at characterizing the nature of such co-locations between two (or more) people rather than an individual’s mobility. Indeed, by examining the frequency of interactions between two people across multiple POIs, we can capture the strength of social forces [15] that result in recurrent co-location of individuals.

### B. Epidemic Modeling

Similar to mobility, epidemics modeling has been largely covered at the micro and macroscale. For instance, at the interpersonal level, researchers investigate avenues of transmission from one person to another by analyzing air droplet trajectories from coughs [16], indoor respiratory transmission [17], and viral durability on different surfaces [18]. Although this vein of research is necessary to identify the mechanisms of transmission at the individual level, it falls short of being able to rule out who could have infected whom at the meso-scale and therefore must be expanded to meet the demand of largescale disease contact tracing.

Conversely, at the macroscale, epidemiologists utilize generalized mathematical and mobility models to simulate diffusive processes related to the disease in question [19]. In the early days of the SARS-CoV-2 outbreak, epidemiologists turned to mathematical models used in 2003 SARS and 2012 MERS outbreak to get an initial estimation of the novel coronavirus’s impact [20]. Though useful in predicting macroscale reproduction numbers, these models are too ambiguous when used at local and city levels. When reflecting on the uncertainty of early predictions, researchers critique that even data-driven mechanistic models are subject to differences in model calibration, data, and assumptions that may lead to contradicting information [21,22].

In response to this critique, a confluence of computer scientists and epidemiologists opted for a data-driven approach that addresses the heterogenous mixing of populations to estimate a spatial outbreak. Some researchers chose to use census data [23], commuter trajectory surveys [24], and mobility tracing datasets such as SafeGraph [25] to estimate interactions. Though these data sources provide an insight into the population’s general mobility, these works start from real data and then generate synthetic interactions which contributes to large estimation uncertainties.

With FS data being available, rather than simulating population mixing, we have access to individual visits and dwell times which can serve as a proxy for digital contact tracing. Our use of the FS POI dataset and mechanistic model maintains the granularity of interactions between individuals, while being scalable enough to apply to large populations. We build on the prior work on both mobility and epidemics to 1) analyze the strength of co-mobility among a city’s population whose colocations then 2) serve as potential avenues for disease transmission.

## III. Approach

In this section, we propose a novel network-based epidemics surveillance method to 1) sense meaningful interactions from real mobility data and 2) predict disease transmission at the meso-scale. To support our investigations, we utilize the FS dataset that contains over 1.3 million individuals and their respective POI visits in Austin, Texas, throughout 2019 and 2020.

To this end, we first build dynamic contact networks where a node is a person, and an edge is formed when two people co-locate at a POI for over 15 minutes. Second, we deploy a classic Susceptible-Exposed-Infected-Recovered (SEIR) epidemic network diffusion model [26] and fit model calibrations to Austin’s COVID-19 infection dynamics. Third, we explore whether we can better our predictions and reduce model uncertainty by pruning the networks for stable interactions. Finally, we verify our approach’s scalability by extending our analysis to NYC.

The strength of the FS dataset differs from existing competitors in that it contains visits, dwell times, and demographic information about each user such as age, home zip code, and sex. This granularity exceeds the usefulness of, say, SafeGraph’s aggregated POI traffic by giving us explicit visit information at the individual level. Furthermore, compared to collecting raw GPS trajectories from sources like X-mode [27], FS employs stop detection to give usable dwell times for each visit. To preserve privacy, FS assigns anonymous identifiers and filters out all predicted home and work visits. Though it would be helpful to know someone’s living situation or co-worker cohort for digital based contact tracing purposes, these POI visits capture when someone makes a non-work-related trip outside of home and can therefore give us some insight into the *social* gatherings between two or more individuals.

Though these largescale dynamic contact networks offer a network topology through which we can simulate who infects whom, let us consider the potential sources of model uncertainty exemplified in Figure 1. In each day of the SEIR simulation, a contagious person infects a percentage of its neighbors. We tune the percentage of infected neighbors when fitting our model to the real infection curves in Austin and NYC. Furthermore, we set the incubation period to 5 days and the infectious period to 12 days to reflect COVID-19 virality as supported by [28].

**Figure 1.**
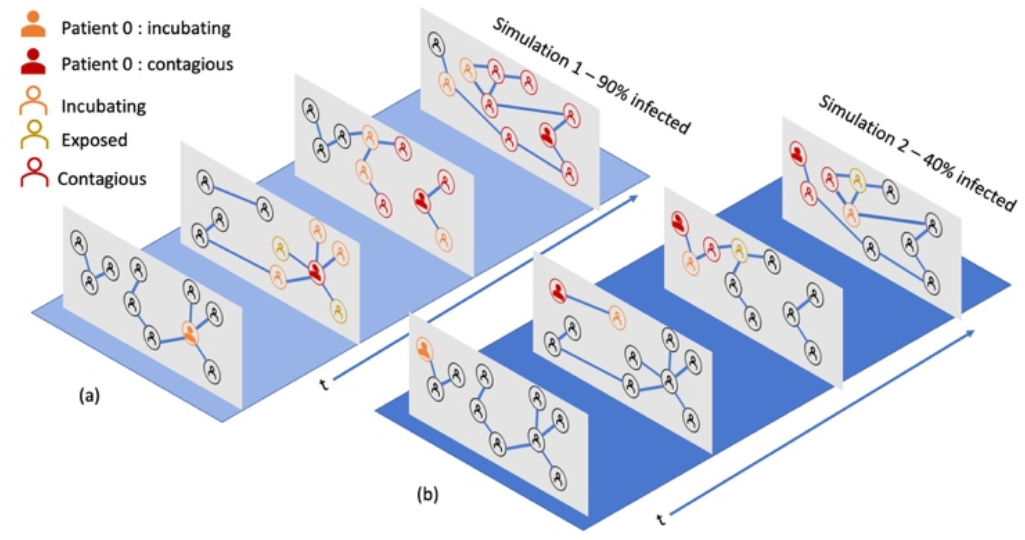
Example of two SEIR simulations run on the same dynamic contact networks varying only patient zero (shown in red). By the end of simulation in scenario (a), 90% of nodes are either infected or incubating (red or orange) versus the 40% of nodes being infected or incubating in scenario (b).

As shown in Figure 1, the diffusion process is inherently dynamic because of time evolving node transitions, as well as the dynamic edges. We can see that the simulation outcomes vary greatly by virtue of who is patient zero. In this toy example, the two simulations result in predictions ranging from 90% of nodes infected (1a) to less than half (1b). The wide range of predictions could make for unreliable interpretations of a city’s outbreak threat level and therefore need to be addressed when performing meso-scale epidemiological analysis.

Upon this observation, we hypothesize that our network-based COVID-19 simulations would be more accurate if we sense recurring interactions that are likely to serve as disease vectors in the real world, and then use those as the backbone topology to simulate our SEIR epidemiological diffusion process. Though the POI in FS data do not allow us to know whether the members of these communities know each other personally, their frequency of interaction can indicate the higher probability of being in each other’s presence on a recurring basis.

Consider the ego network shown in Figure 2a where the central person (in grey) interacts with those on the periphery through different contexts and frequencies. Although we do not explicitly know the social relationship behind their interactions, we do know the type of POI (e.g. coffee shop, gym) and how often these people co-locate.

**Figure 2.**
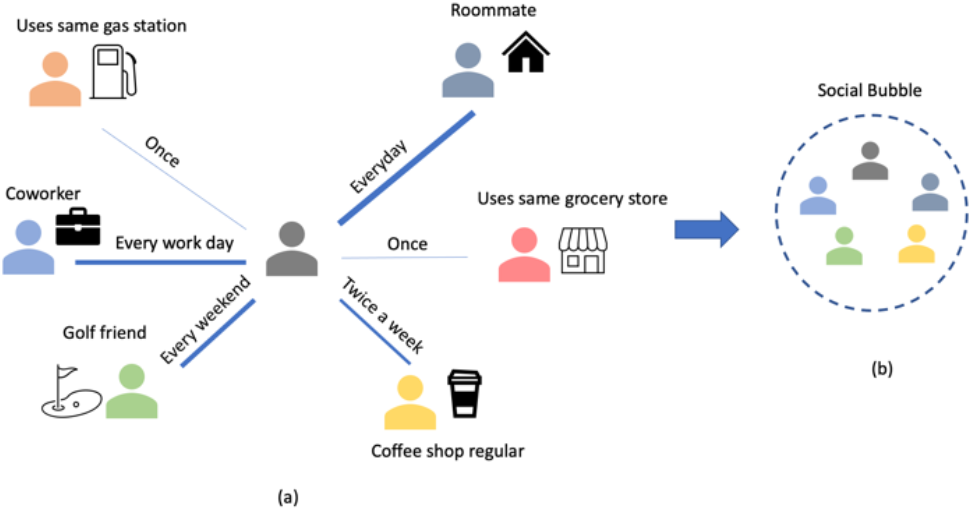
(a) Ego network exemplifying various strengths of repeated co-locations. By studying frequency of interaction and context of the Point of Interest (POI), we can get a sense of the nature of predictability behind these interactions that lead to forming the social bubble shown in part (b).

For example, let’s say that the orange person filled up the gas tank near the grey person long enough to show up on our co-location network. However, this interaction is not likely to repeat, nor lead to a real infection; therefore, the edge between these two people could be safely removed from the simulation space. Similarly, the person in pink frequents the same grocery store as the dark grey person but has only co-located with them once. In contrast, the frequency of interaction with the light grey, blue, green, and yellow people indicate a regularity that could more likely propagate a virus among the group. The interactions in part 2a can be abstracted to the social bubble shown in Figure 2b where from the perspective of a highly transmissible disease, these reliable interactions can serve as a point of vulnerability if one of these people contracted the virus.

Let us now revisit the toy example discussed in Figure 1, however this time using pruned networks shown in Figure 3a and 3b. As before, the only variation between simulation runs is the choice of patient zero. Although the runs result in both different paths and overall number of infections, the spread of uncertainty decreased substantially where the simulation in Figure 3a has 50% of nodes infected while the simulation in Figure 3b has 40%. This reduction of noise is intuitive because the traversal space for the infection is limited to reliable interactions rather than insignificant interactions.

**Figure 3.**
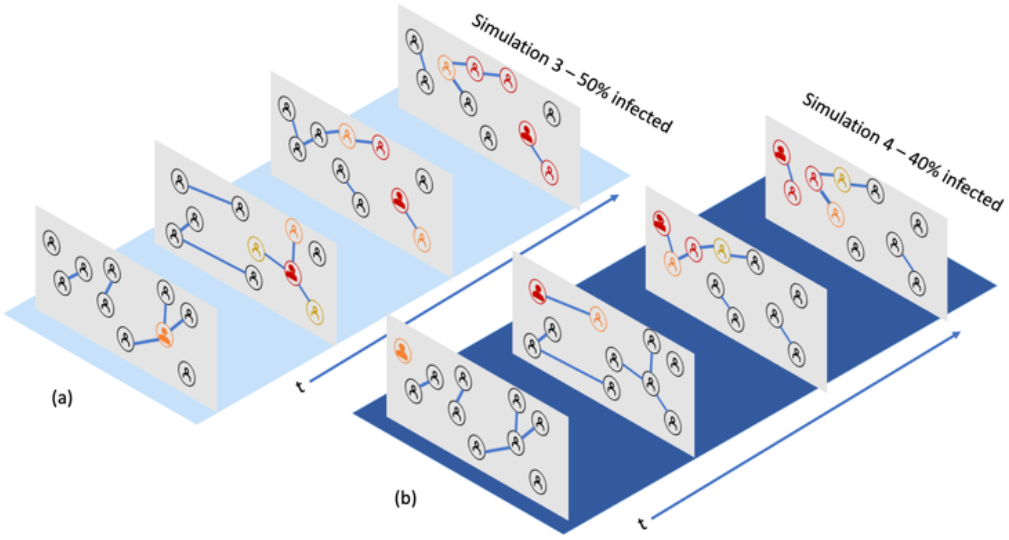
Rerunning SEIR in Figure 1 on a pruned dynamic contact network results in reducing the model uncertainty between simulation 3 (a) 50% of nodes infected and simulation 4 (b) 40% infected. This shows the importance of pruning contact networks for stable interactions.

We define our model as such: let *G*_*t*_ = (*V*_*t*_, *E*_*t*_) represent one co-location network at time *t* where *V*_*t*_ is the set of nodes and *E*_*t*_ is the set of edges. We define the *aggregated frequency network as F =* (*P, R*) where *P* are the set of nodes and *R* are the set of weighted edges. We build *F* by iterating over *N* co-location networks *G* = [*G*_1_, *G*_2_, … *G*_*N*_]and aggregate the edges as shown in the equations below.

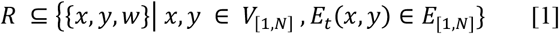

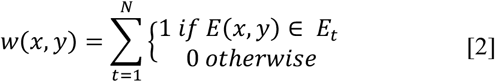

To calculate the edge weight *w*(*x,y*), we keep a counter of how many times an edge *E*_*t*_ (*x,y*) occurred in the *N* co-location networks as shown in equation 2. For example, if an edge *E* (*x,y*) ∈ [*E*_1_, *E*_4_, *E*_8_] then *W*(*x,y*) =3. After forming the frequency network *F*, we prune the original co-location networks *G* to contain the edges that have a weight of λ or more, where λ is the minimum frequency of interactions within a period that we deem to be non-volatile. Rather than using the raw co-location networks that considers when two people have been near each other only once, we hypothesize that pruning dynamic contact networks for stable connections decreases the noise and reveals more accurate results.

In summary, our approach first builds co-location networks from FS data, then aggregates the networks to see which interactions recur vs those that are fleeting. Once volatile edges are identified, we prune them from the co-location networks leaving the network topology of the recurring social bubbles. We hypothesize that agent-based modeling on the pruned co-location networks will yield less noisy and more accurate results. As a departure from the state of the art, our approach allows for the confluence of digital contact tracing and epidemic predictions at the meso-level.

## IV. Results

We first perform general contact network analysis on Austin, TX networks to verify the expected trends resulting from the COVID-19 lockdowns. After exploring the metrics, we then deploy our pruning and mechanistic model on both Austin and NYC contact networks.

When first comparing Austin’s network metrics (e.g., number of nodes, average degree, and clustering coefficient) for both 2019 and 2020, we notice a major discrepancy in the volume of data. To investigate the discrepancy, we note the raw number of POIs visited per day as shown in Figure 4. As expected, the number of POIs visited substantially drops from 2019 to 2020 even before SARS-CoV-2 reached Austin in mid-March. Upon further investigation, we find this trend to hold across the entire FS dataset. To account for the underlying volume difference in data between these two years, we normalize the number of nodes (Figure 5) based on the population percentage rather than raw number of people.

**Figure 4.**
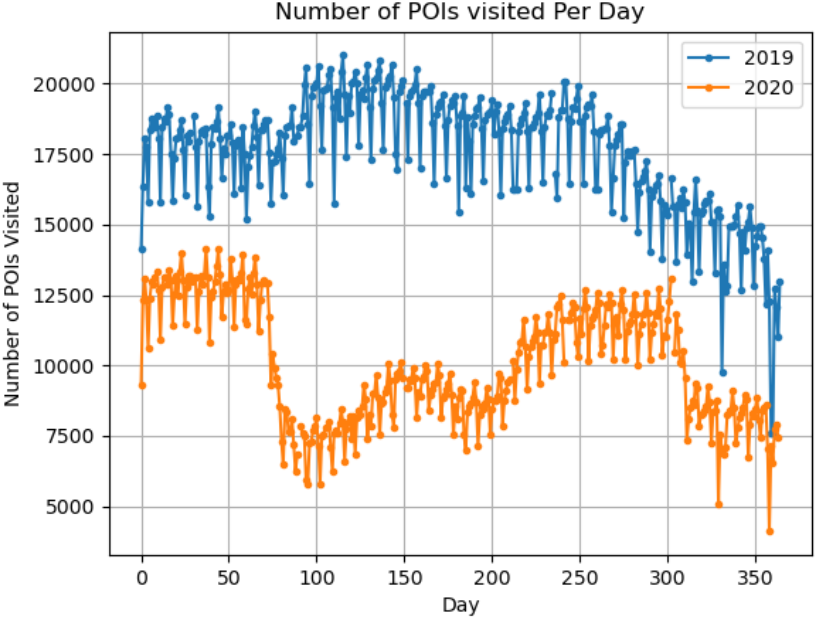
Comparison of Points of Interest (POI) visited per day in Austin Texas for years 2019 and 2020. Though, the Austin COVID-19 lockdown occurred on day 75 (March 15^th^), the raw POI visits were less than in 2019 even before the lockdown.

**Figure 5.**
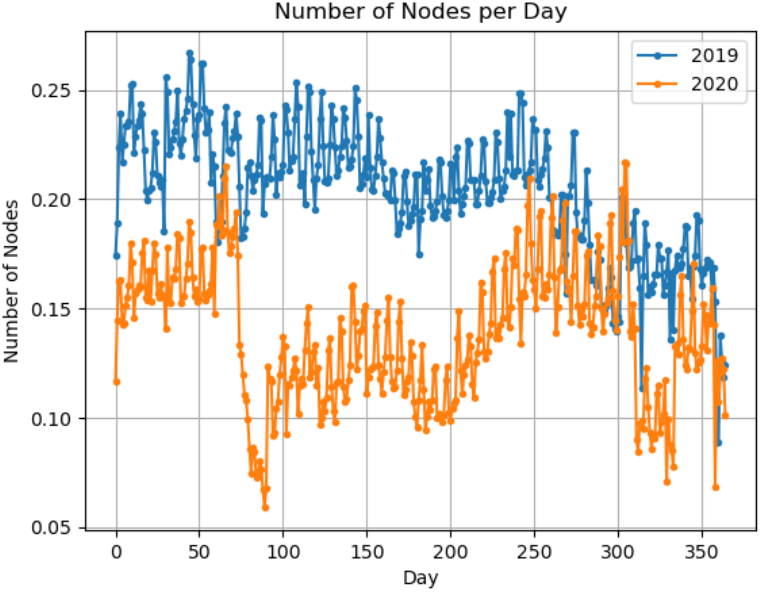
Number of nodes in co-location networks per day from 2019 through 2020 in Austin TX. To account for the population difference, we show the normalized number of nodes that makeup the subset of people co-locating at POIs.

### A. Co-location Network Analysis

As shown in Figure 5, the number of people co-locating at POIs fluctuate according to Austin’s lockdown and lifting restrictions. The 2020 co-location networks’ most dramatic deviation from 2019 is on March 15^th^ (day 75) when Austin shuts down all non-essential businesses and mandates social distancing. The number of co-locations then later increases in response to the relaxation of lockdown orders in the summer of 2020. Furthermore, the minor fluctuations on a smaller scale show that the size of co-location networks correspond to the day of the week where larger congregations of people take place over the weekend versus during the weekday.

Similarly, Figure 6 shows that the average node degree in 2019 and pre-pandemic 2020 fluctuates with the day of the week where most people have more co-locations on the weekend than during the weekday. This makes sense because the FS data only records non-work and non-home POI visits. Furthermore, the 2019 major blips correspond to Austin’s music festivals in the spring, summer, and fall. In response to COVID-19 lockdown on March 15, 2020, the average degree declines reflecting the effort to minimize non-essential trips and socially distance. The decline in number of nodes per day indicate an overall decrease in the population co-locations whereas the decline of average degree per day shows that those who are co-locating are encountering fewer people on average compared pre-pandemic POI behavior.

**Figure 6.**
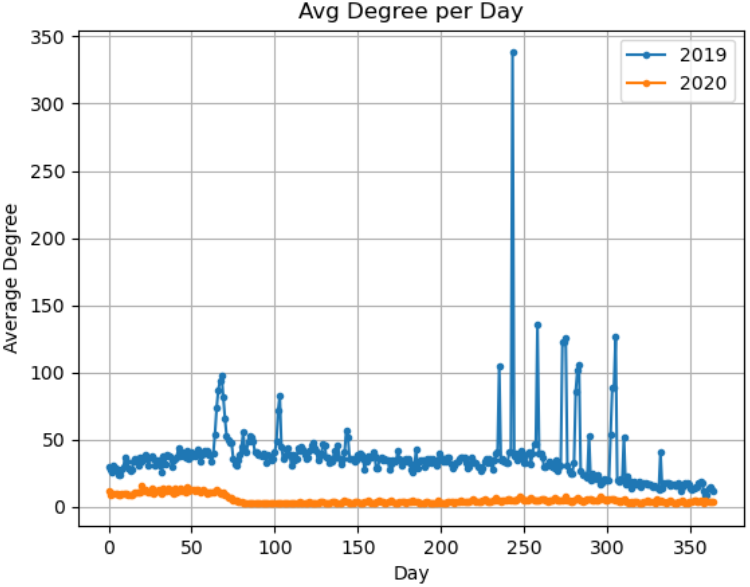
Average node degree in co-location networks per day from 2019 through 2020 in Austin TX. The largest peak in 2019’s average node degree corresponds to a major music festival which connects many people co-located at the concerts.

In Figure 7, the clustering coefficient fluctuations reveal Austin’s *volatility in connectivity* as less people are visiting POIs at the same time. In addition, we note that Austin’s co-location networks became more disjointed in comparison to the 2019 networks. This suggests that people within Austin, whether through city mandate or personal intention, were adhering to forming social interaction bubbles.

**Figure 7.**
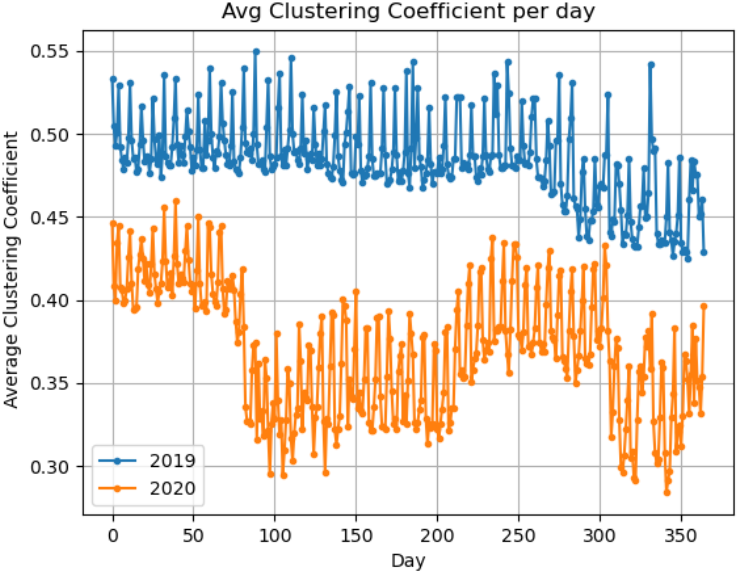
Average clustering coefficient in co-location networks from 2019 through 2020 in Austin Texas. As shown above, Austin’s connectivity fluctuates in response to COVID-19 lockdown restrictions making the city more disjointed as a means to mitigate disease transmission.

### B. Pruning Networks

In effort to sense the COVID-19 social bubbles that formed in response to the lockdowns, we aggregate the co-location networks within a month to capture the frequency of stable interactions. We then validate the significance of the interaction frequency by building synthetic random networks from the original population and then shuffling their connections. For example, if a pair of nodes have a co-location frequency of 14 times within one month period, their nodes would get 14 edge rewires in the synthetic network. By shuffling the links, we test whether a frequency occurs by chance or is statistically significant. We then compare the co-location frequency in Figure 8, which reveals a large gap between the real frequency networks and the random control networks. Furthermore, the largest amount of people with higher co-location frequencies occurred for both Austin (Figure 8a) and NYC (Figure 8b) in January of 2020, compared to the post-pandemic co-location networks. Additionally, as shown in the ‘Random Interaction’ curves in Figure 8(a-b), the percentage of population that randomly co-locates more than twice a month is less than 1% during the pandemic and under 3% pre-pandemic. This informs our decision to prune the edges that only occur once within a month from our dynamic contact networks.

**Figure 8.**
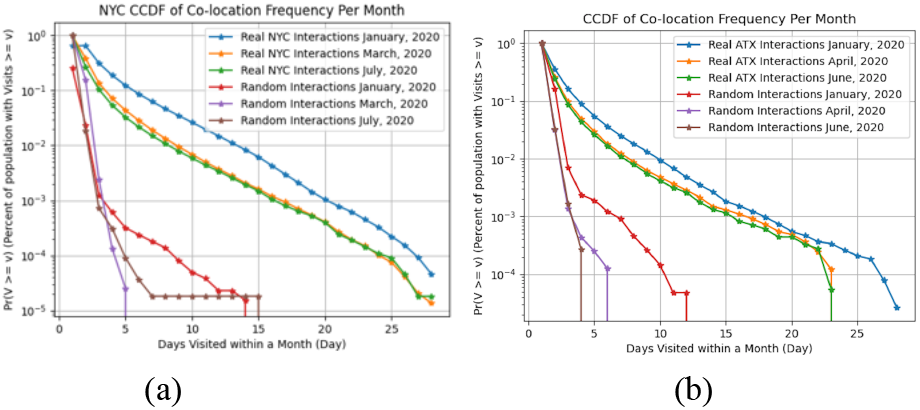
Complimentary Cumulative Distribution Function (CCDF) comparing the frequency of visits throughout Austin (a) and NYC (b) for pre-pandemic and post-pandemic months. The CCDF shows both the discrepancy in real frequencies vs synthetic frequencies, as well as the decrease in co-location frequency due to lockdown restrictions.

### C. Modeling COVID-19 Analysis

After building both raw co-location and pruned networks, we deploy the SEIR diffusion model on both sets of networks to compare against the real infection outbreak recorded during July 2020 in both Austin and NYC. We hypothesize that the pruned networks will not only reveal a narrower set of potential outcomes, but also provide more accurate predicted number of infections per day.

As shown in Figure 9(a-b), the SEIR simulations based on raw co-location contact networks (in red) overshoot the real infection curve (in green) exponentially. This can intuitively be explained by the concept previously pointed out (see Figure 1) where volatile interactions could account for the difference between simulations diffusing the infection across network hubs and bridges. In essence, the infection can traverse any interaction and thus amplify the “noise” stemming from the volatile interactions.

**Figure 9.**
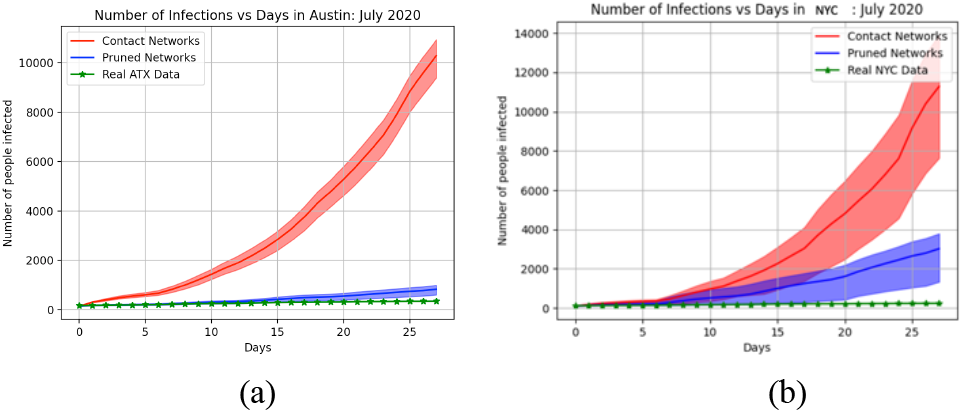
Network-based SEIR simulation runs on raw dynamic co-location networks (red) and pruned dynamic co-location networks (blue) for the month of July 2020. Both real Austin (a) and NYC’s (b) reveal a closer fit to the simulation runs when using pruned networks.

In comparison, the pruned interactions reveal a much more reasonable fit to Austin’s real infection outbreak going from an average residual of ±3,163 people to only ±140 people. Similarly, simulating an outbreak on NYC’s pruned interactions decreased the average residual by 70%.

To account for the effect of volatile interactions on the diffusion process, we vary the percentage of noise (i.e., co-locations that occur only once within a month) preserved in the pruned networks. As hypothesized, the volatile interactions accelerate the disease process and thus amplify the noise in a characterizable way (Figure 10).

**Figure 10.**
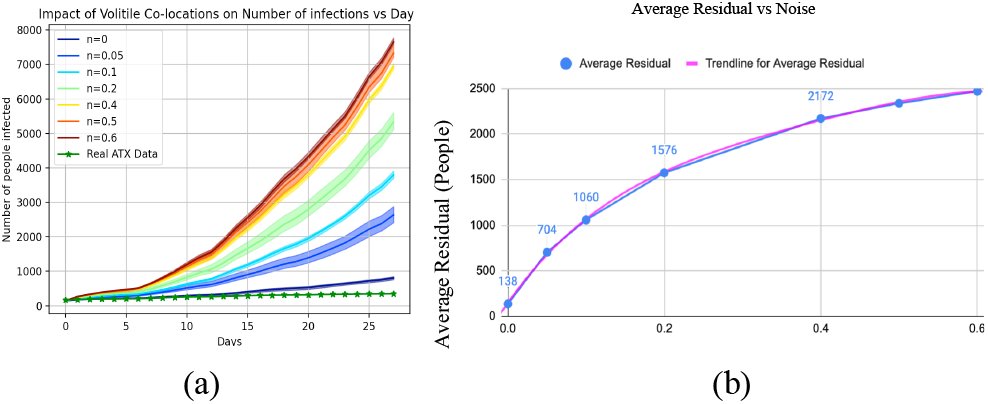
Average residual from simulated data with Noise (Percent of volatile edges) used in simulating SEIR through pruned co-location networks. When comparing the curve of real infections to the simulations, the insertion of volatile interactions exhibits a polynomial relationship to the average error.

The average residual vs percent of preserved volatile edges depicted in Figure 10 show a polynomial relationship that emphasizes the amplification of noise. This can be interpreted intuitively as in all the simulation runs, namely a disease can traverse through a network in a plethora of ways. Every volatile interaction gives the simulation run an option to spread the virus to a new region in Austin or NYC, or a new social bubble. Though a volatile interaction may be the fault of a community catching a transmissible disease, when simulating at a city-level, the noise of these interactions overpowers the quality of information gained from these digital mobility-based epidemic simulations.

Epidemic predictions become useful to policy makers when they can be used to inform not only the size of the outbreak but also the regional severity of a forecasted outbreak. The map in Figure 11 (a) shows a normalized view of the July 2020 spatial outbreak of COVID-19 Austin. Figure 11 (b) depicts the forecasted spatial outbreak from the pruned interactions, and lastly the map in Figure 11 (c) shows the forecast from raw co-locations. Figure 11(e-g) exhibit the same comparison between real cases (e), simulate cases from pruned interactions (b), and all interactions (g) in NYC. As in the case for both Austin and NYC, simulating an agent-based epidemic model using digitally traced interactions leads to an exaggeration of threat level both on the number of infections predicted, as well as the spatial representation of the outbreak.

**Figure 11 (a-c).**
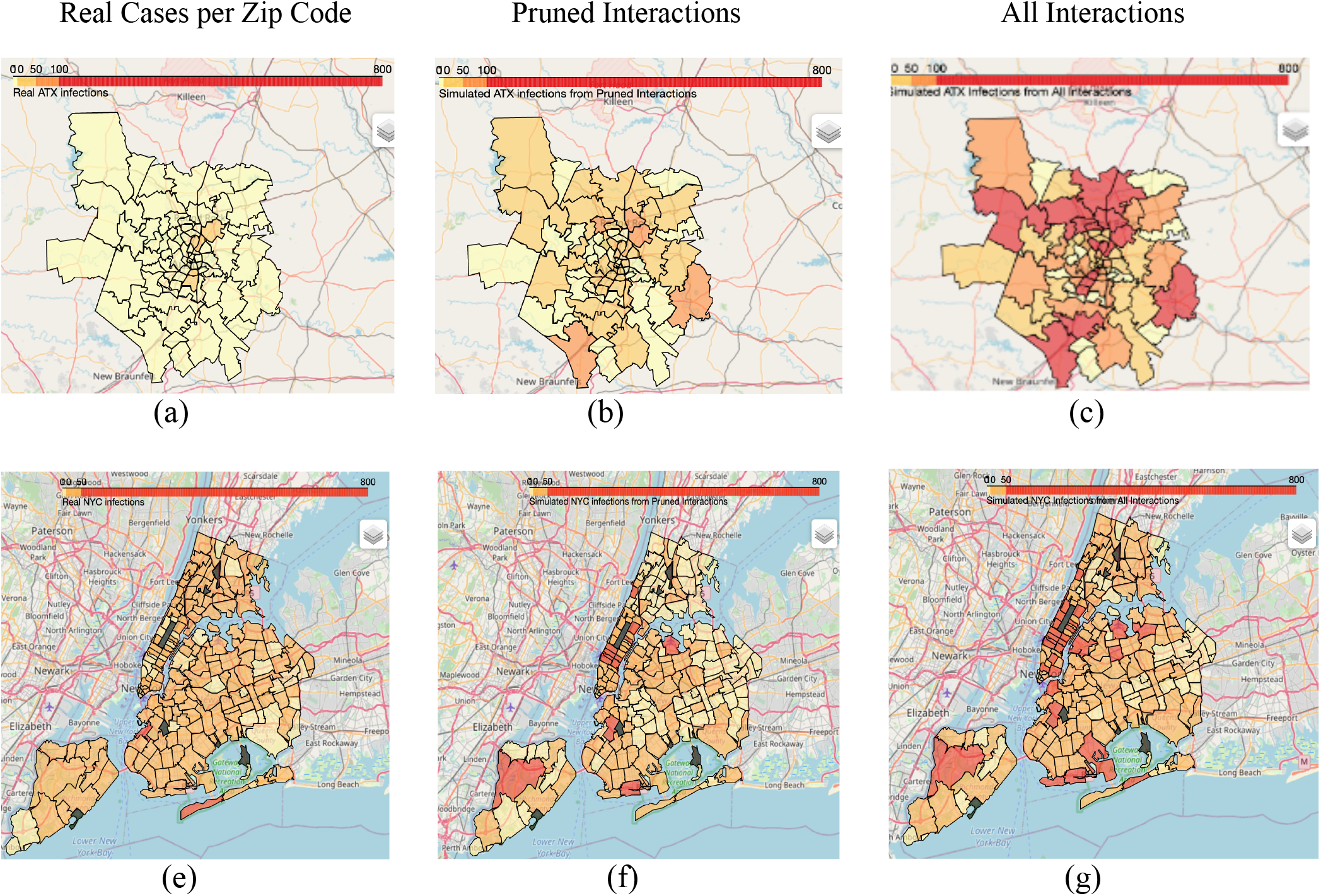
Comparison of real COVID-19 infection spread in Austin during July 2020 (a) and the network-based SEIR simulations using (b) pruned interactions and (c) all interactions. The maps shown in (e-g) exhibit the same comparison using NYC real cases (e), simulated on pruned interactions (f), and on all interactions (g). The dark grey regions in NYC correspond to city park zipcodes that do not have residents.

## V. Conclusion and Future Work

In this paper, we have proposed the use of smart-phone enabled mobility tracking and network-based methods to both *scale up* contact tracing and *scale down* epidemic modeling to provide actionable disease prediction at the meso-scale. Using real interactions taking place at POIs throughout Austin and NYC, we have modeled outbreaks directly from digital contact networks to get an estimate of not only of their size, or which region, is getting sick –but also identify the individuals likely to get infected.

We have found that directly applying the classic SEIR model to dynamic contact networks can result in large uncertainty due to volatile interactions. In the mobility data, we have observed and utilized the fact that people regularly co-locate among different POIs revealing groups of people likely to infect each other if contagious. Furthermore, we have found that the co-location frequencies reveal a power law distribution; we can interpret the range of co-location frequencies as the strength of attraction between two people that then form stable micro-communities within the city.

We have found that pruning dynamic contact networks for stably reoccurring interactions gives a more accurate city-level topology for which an infectious disease is likely to traverse. When compared to real number of infections vs time, our pruning method decreases the average error of simulated number of infections from ±3,163 people to ± 140 people in Austin’s projections and reduce noise by 70% in NYC’s simulations. Our method to prune dynamic contact networks not only provides a solution to denoise large spread of city-level health outcomes, but also lays the groundwork for a scalable, fast-acting, ‘always on’ approach to epidemic surveillance.

Future work can extend our noise reduction method by installing a feedback loop to allow people to indicate whether they have the virus. Both reducing the volatile interactions and updating which people are sick during the simulations can lead to a framework that simultaneously digitally contact traces and monitors an ongoing outbreak at the meso-scale. Building such framework will serve as a major contribution in the effort to plan for the next pandemic.

## Data Availability

Data is privately sourced from FourSquare made available through AWS.

